# Mendelian randomization study on the association between cheese intake and diabetes

**DOI:** 10.1101/2024.01.12.24301232

**Authors:** Si-Qi Yang, Yu Xu, Yan-Bing Zhang, Wei-Xing Wang

## Abstract

**Objective:** To explore whether there is a causal relationship between cheese intake and diabetes through Mendel randomization (MR).

**Methods:** Two samples of MR were used to verify the causal effect of cheese intake on diabetes. The analysis was conducted using inverse variance weighted (IVW), weighted median, and MR-Egger regression methods. We used a meta-analysis of publicly available genome-wide association studies (GWAS) to compile statistical datasets, and cheese intake data as exposure factors were sourced from individuals of European ancestry in the UK biobank (n=451486). At the same time, GWAS’s public summary statistical data set is also used for self reporting non cancer disease codes: diabetes data included in the Finnish database (total n=184404; case=1219, control=183185) (http://www.finngen.fi/en)as a result.

**Result:** We selected 108 single nucleotide polymorphisms (SNPs) with genome-wide significance as instrumental variables from the GWAS intake of cheese. IVW method results show a causal relationship between cheese intake and diabetes (β=- 1.196, SE=0.3817, P=0.001729). MR Egger regression results show that directed pleiotropy is unlikely to bias the results (intercept=0.015; P=0.58), but there is no causal relationship between cheese intake and diabetes (β=- 2.073, SE=1.621, P=0.206). The results of weighted median method also showed that there was no causal relationship between cheese intake and diabetes (β=- 0.7828, SE=0.5701, P=0.1698). Cochran’s Q-test and funnel plot indicate no evidence of heterogeneity and asymmetry, indicating the absence of directed pleiotropy.

**Conclusion:** The results of MR analysis support that cheese intake may have a causal relationship with the reduced risk of diabetes.

## 1. Introduction

Diabetes is one of the common chronic diseases affected by many factors, including diet patterns, weight gain and lack of exercise can promote the onset of diabetes^[1-3]^. In 2021, the number of adult diabetes patients in the world will reach 537 million^[4]^. Some studies predict that by 2050, this number will more than double, reaching about 1.3 billion people, with a incidence rate of nearly 10%, far exceeding the current incidence rate of 6%^[5]^. The progress of diabetes will also lead to other related complications, which will bring major obstacles to social, economic, medical and other development^[6-8]^. The chronic complications of diabetes involve large and small blood vessels of the whole body, and the pathological changes extensively involve many organs such as the heart, kidney, nerve, etc., which is the main reason for the decline of patients’ quality of life and death. Microangiopathy is a direct consequence of hyperglycemia and a specific complication of diabetes. Its occurrence is related to the course of disease and the degree of hyperglycemia. Therefore, controlling blood sugar can effectively reduce the complications related to eye, kidney and neuropathy caused by diabetes.

In particular, the onset of type 2 diabetes is insidious, nearly 50% of diabetes patients have no obvious symptoms, and the potential complications are difficult to detect, which often attract attention when there are symptoms such as significant decline in vision, renal failure, limb numbness and pain, thus losing the opportunity for effective treatment. The prevalence of retinopathy and microalbuminuria in individuals with impaired glucose tolerance (IGT) is as high as 10% and 16%, which are 3-4 times higher than the normal glucose tolerance population. Atherosclerosis is a common macrovascular complication of diabetes, which can lead to coronary heart disease, cerebrovascular disease and peripheral vascular disease. Foreign epidemiological studies have confirmed that 50% of diabetes patients are accompanied by atherosclerosis, and the mortality rate after the first myocardial infarction is significantly increased, which is equivalent to the mortality rate of coronary heart disease patients without diabetes after the second myocardial infarction^[8]^. Atherosclerosis associated with hyperglycemia already exists in IGT stage. A study by Fungata in Japan found that IGT patients diagnosed based on 2-hour postprandial blood glucose have an increased mortality rate, while patients with impaired fasting glucose (IFG) have a similar mortality rate to those with normal fasting blood glucose^[9]^. The 5-year follow-up data of diabetes and cardiovascular and cerebrovascular events risk in China showed that the risk of cardiovascular and cerebrovascular events in diabetes patients increased 4.78 times and 3.78 times respectively, and the risk of cardiovascular and cerebrovascular events in IGT patients also increased 3 times^[10]^. Compared with the treatment after illness, how to reduce the incidence rate of diabetes and prevent the further development of diabetes by adjusting the diet structure has become a concern of more and more clinicians.

A large number of meta-analysis results have shown that improving the overall diet quality can significantly reduce the incidence of diabetes, especially type 2 diabetes (T2D)^[11-12]^. Among them, the intake of dairy products is of great concern, because dairy products contain rich minerals such as calcium and magnesium, which can significantly reduce the risk of diabetes and insulin resistance^[13]^. In addition, whey protein has proinsulin and hypoglycemic properties by secreting insulin and intestinal growth hormone^[14]^. In recent years, the correlation between dairy products and the risk of diabetes is controversial. Meta analysis results show that the consumption of total dairy products is inversely proportional to the incidence of diabetes, especially the consumption of yogurt, but not all research results are consistent^[15]^. As a fermented dairy product, cheese is rich in protein, calcium, saturated fatty acids and other nutrients^[16]^. At present, there is no research to analyze and confirm whether there is a correlation between cheese intake and diabetes.

In recent years, Mendelian randomization (MR) analysis has been widely used to assess the potential causal relationship between various risk exposures and clinical outcomes^[17-19]^. Compared to traditional observational studies, MR analysis can overcome reverse causal bias due to allele randomization occurring before disease onset^[20]^. In addition, the random separation and independent classification of genetic polymorphisms during conception enable MR analysis to minimize the impact of confounding factors by introducing genetic markers as instrumental variables (IV) for exposure. The availability of large-scale genome-wide association studies (GWAS) further enables the exploration of causal relationships^[21]^. Therefore, from the overall perspective, this paper explores the positive correlation, negative correlation or no causal relationship between cheese intake and diabetes (no complications) by applying MR analysis.

## 2. Data source

The MR Base database(http://www.mrbase.org/) contains a large collection of statistical data from hundreds of genome-wide association studies (GWAS). We searched using the publicly available GWAS meta-analysis summary statistical dataset, analyzing cheese intake data of European descent (n=451486) as exposure factors. We screened GWAS data to include SNPs significantly associated with cheese intake. Based on the results of the cheese intake meta-analysis, SNPs with a correlation of P<5E-08 were included as instrumental variables(IVs); And in order to avoid the impact of linkage imbalance (LD) in SNP on the analysis results, the parameter r^2^ threshold is set to 0.001 and the distance is set to 10000 kb. In order to ensure a strong correlation between instrumental variables and endogenous variables and avoid weak instrumental bias, we calculated r^2^ for each SNP, representing the proportion of variance explained by the instrumental variable SNP; The F-statistic is used to evaluate the strength of the instrumental variable based on a P-value threshold of 5.00E-08 (whole genome significance). Genetic variations related to cheese intake were screened as IVs, and two sample MR analysis was performed. We have screened a total of 108 single nucleotide polymorphisms (SNPs) related to cheese intake and summarized statistical data(β Coefficient and standard error) as IVs. We used data on uncomplicated diabetes published in the Finnish database(total n=184404; case=1219, control=183185)(http://www.finngen.fi/en)as a result.

## 3. Mendelian randomization analysis data

Mendelian randomization is the use of genetic variation in non experimental data to estimate the causal relationship between exposure and outcome^[22]^. After evaluating the independent correlation between SNPs and cheese intake, we studied the relationship between each SNP and the risk of diabetes. Next, we combined the above findings and used MR analysis to assess the correlation between cheese intake and the risk of diabetes. Due to the need to estimate the causal relationship between exposure factors (cheese intake) and results (diabetes), we selected two samples of MR. We chose to use the summary statistical data of different GWAS to evaluate the causal relationship between cheese intake and diabetes, and finally selected 108 SNPs as IVs (Table 1).

**Table 1.** The causal relationship between cheese intake and diabetes risk assessed by each MR analysis method.

| MR method | Number of SNPs | Beta | SE | 95% confidence interval | Association P-value | Cochran Q statistic | I<sup>2</sup> | Heterogeneity P-value |
| --- | --- | --- | --- | --- | --- | --- | --- | --- |
| Inverse-variance weighted | 62 | -1.196 | 0.3817 | -1.2558~-1.1362 | 0.001729 | 57.85 | 0.018 | 0.5907 |
| MR Egger | 62 | -2.073 | 1.621 | -2.17665~-1.96935 | 0.206 | 57.54 | 0.015 | 0.5661 |
| Weighted median | 62 | -0.7828 | 0.5701 | -0.82194~-0.74366 | 0.1698 | / | / | / |

The inverse variance weighted (IVW) method uses meta-analysis to combine the Wald ratio estimates of causal effects obtained from different SNPs, and provides a consistent estimate of the causal effects of exposure on the results when each genetic variation satisfies the IV hypothesis^[23]^. Its characteristic is that the existence of intercept terms is not taken into account in regression and the reciprocal of the outcome variance (the quadratic power of se) is used as the weight for fitting^[24]^. Although the inclusion of multiple variants in MR analysis increases statistical ability, it is possible to include pleiotropic genetic variants of ineffective IVs. In order to explore and adjust pleiotropy, that is, the association between genetic variation and multiple variables, weighted median and MR Egger regression methods were used. The MR Egger method is widely used in Mendelian randomization analysis that uses multiple SNPs as instrumental variables for causal inference, especially when there is directed pleiotropy in genetic variation^[25]^. The core of this method is to consider the existence of intercept terms in weighted linear regression, which is used to measure the average pleiotropy between instrumental variables, and the slope is an unbiased estimate of causal effects^[26]^. The weighted median (WM) is the median of the distribution function obtained by ranking the SNP effect values of all individuals according to their weights^[27]^. When at least 50% of the information comes from effective instrumental variables, WM can obtain robust estimates^[28]^. Compared with MR Egger, WM has the advantage of higher estimation accuracy. A P-value less than 0.05 is considered statistically significant. All MR analyses were conducted on the MR Base platform (application version 1.2.1 e646be [June 27, 2018], R version 3.5.0).

## 4. Heterogeneity and Sensitivity Testing

We evaluated the heterogeneity between SNPs using Cochran’s Q-statistic and I2 statistic, and if P>0.05, it indicates no significant heterogeneity. At the same time, a “leave-one-out” analysis was conducted to observe whether it had an impact on the analysis results, and a forest map was drawn. If a certain SNP is excluded and P>0.05 is obtained, it is considered that the SNP will not have a significant impact on the results.

## 5. Results

### 5.1 Screening Results of Mendelian Randomization Analysis Tool Variables

We screened 108 independent SNPs from GWAS data on cheese intake as IVs. These are significantly correlated with the whole genome intake of cheese (Table 1, Figure 1). The results showed that 62 out of 108 SNPs were negatively correlated with diabetes, and the results were statistically significant (Table 1, Table S1). 2.7% of the exposure variance (r^2^ statistic) is caused by genetic variation in IVs. A P-value of 5.00E-08 corresponds to a F-statistic value >30 for each variable, and when the F-value is less than 10, it is defined as weak IVs. Therefore, weak instrument bias can be ignored.

**Fig. 1.**
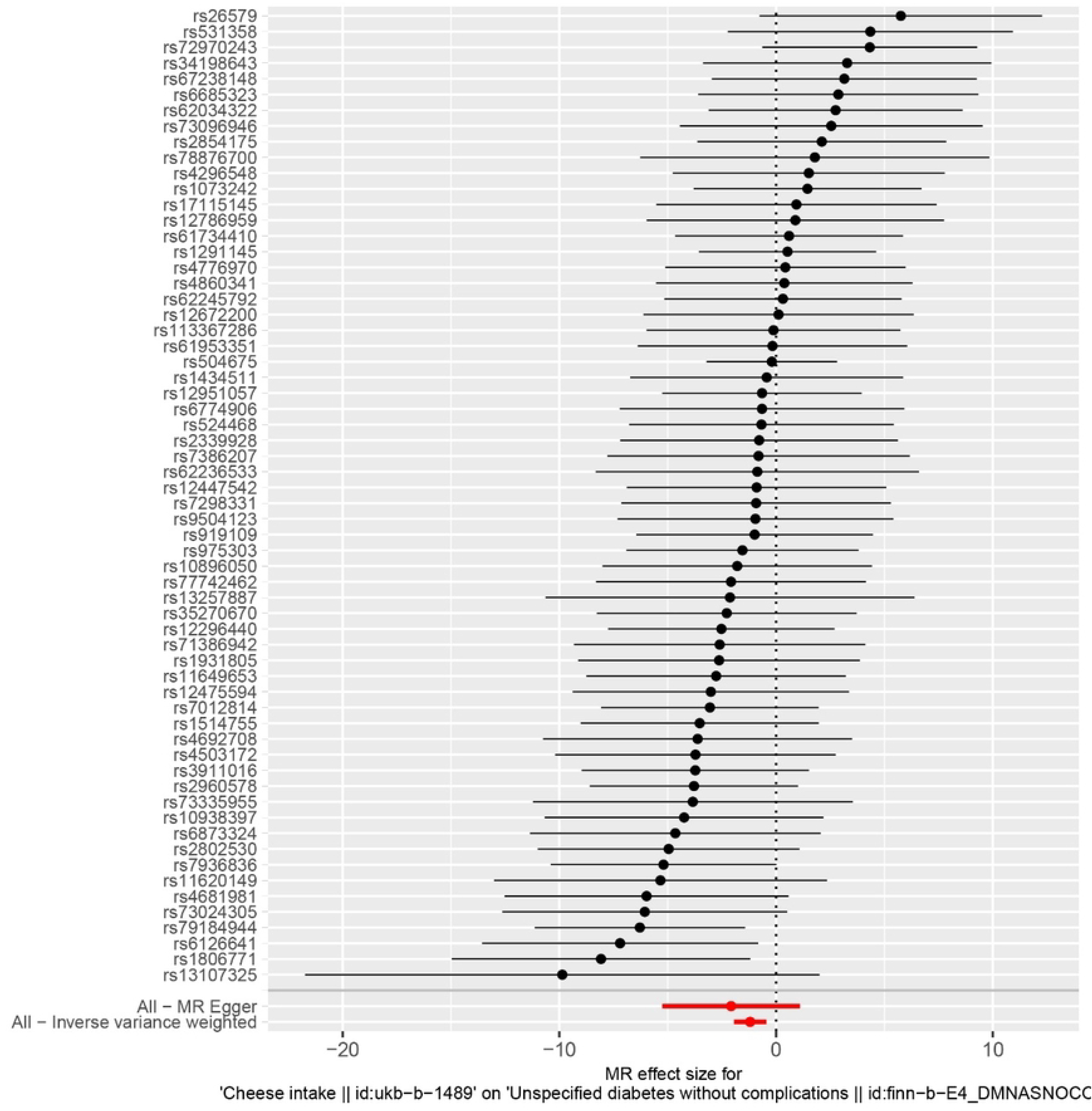
Box initially at rest on sled sliding across ice.

### 5.2 Mendelian Randomization Analysis Results

IVW method results show that there is evidence supporting a causal relationship between cheese intake and diabetes(β=- 1.196, SE=0.3817, P=0.001729; Table 1, Figures 1 and 2). The intercept represents the average pleiotropic effect of genetic variation (the direct impact of variation on the average outcome). If the intercept approaches 0, it can be considered that there is no horizontal pleiotropy.

**Fig. 2.**
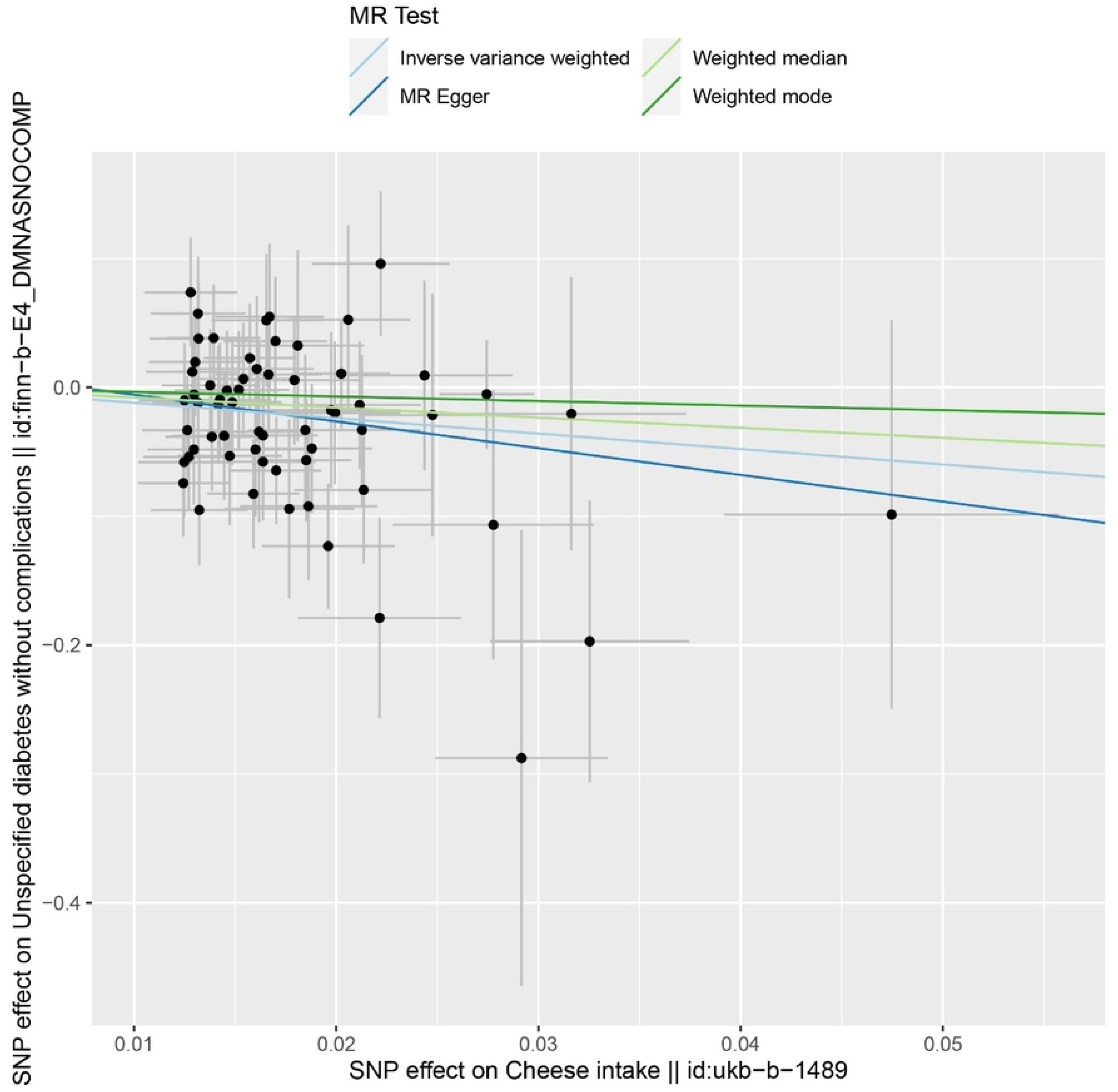
Box initially at rest on sled sliding across ice.

The MR Egger regression results showed that directional pleiotropy had almost no impact on the results (intercept=0.015; P=0.58). MR - Egger analysis shows that there is no causal relationship between cheese intake and diabetes (β=- 2.073, SE=1.621, P=0.206; Table 1, Figures 1 and 2). Similarly, the weighted median method also showed that there was no causal relationship between cheese intake and diabetes (β=- 0.7828, SE=0.5701, P=0.1698; Table 1, Figures 1 and 2). IVW method confirmed that there was a causal relationship between cheese intake and diabetes risk, while MR Egger method and weighted median method showed that the causal effect was zero. Due to the fact that MR is mainly analyzed through the IVW method, which provides the most accurate estimation. Therefore, the results of MR analysis may support the potential causal relationship between cheese intake and diabetes. Due to β value of three methods were all less than 0, indicating that cheese intake may be a favorable factor for the prevention and control of diabetes.

### 5.3 Heterogeneity and Sensitivity Testing

Cochran’s Q-test showed that there was no heterogeneity between IV based on individual variation (Table 1). Heterogeneity refers to the variability of causal estimates obtained for each SNP (i.e., how consistent the causal estimates are for all SNPs). I^2^ value results show low heterogeneity, indicating a high reliability of MR analysis results (Table 1). The results of the “leave-one-out” analysis show that after gradually removing each SNP, the overall error line does not change much (all error lines are on the left side of 0), indicating that the analysis results are relatively stable and reliable (Figure 4). The symmetry of the funnel plot and the intercept of each group in the MR Egger regression test results are equal to or close to 0, and P>0.05 indicates that the results do not have directional level pleiotropy, which also indicates that the MR analysis results are almost unbiased (Figure 3).

**Fig. 3.**
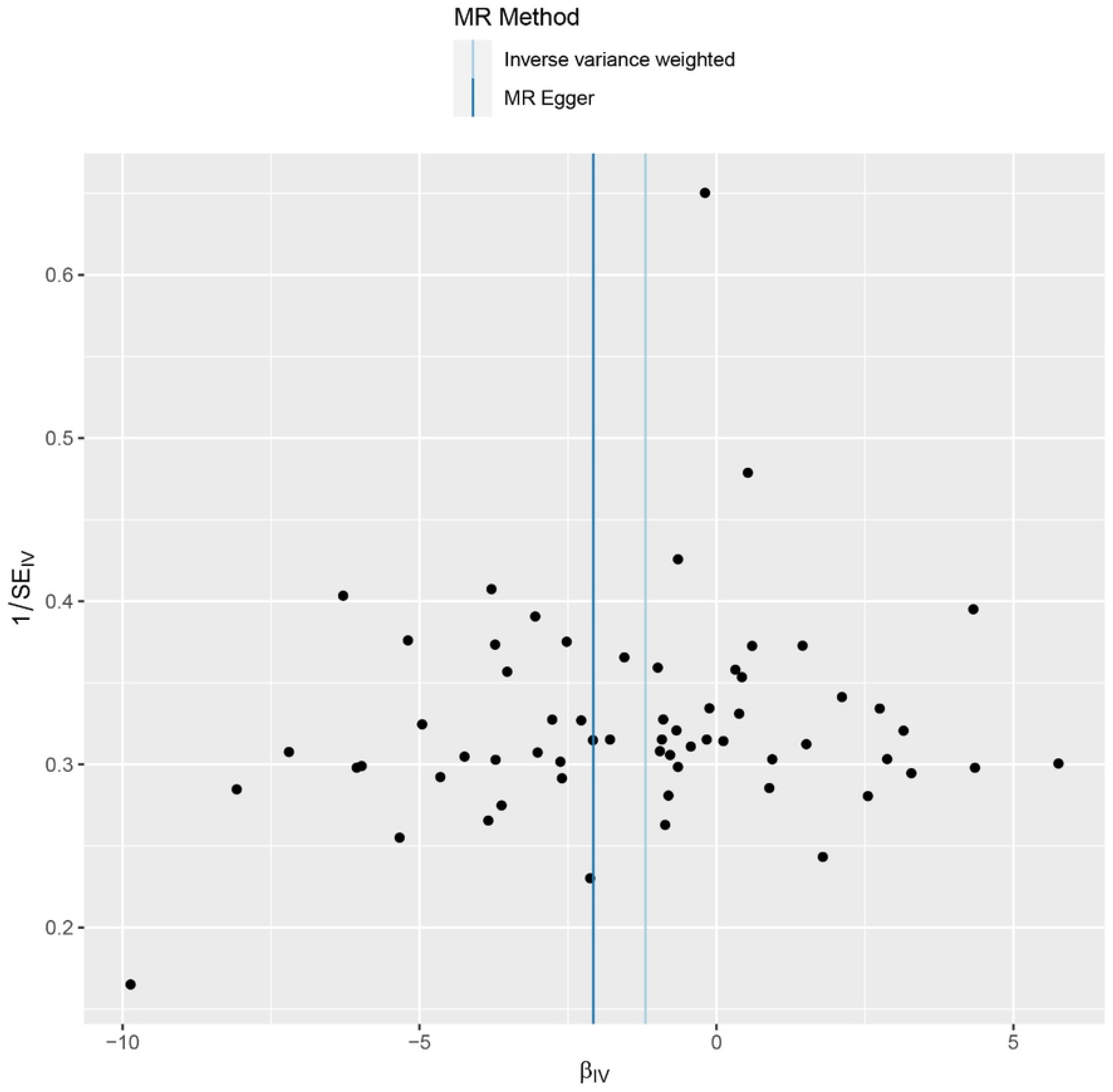
Box initially at rest on sled sliding across ice.

**Fig. 4.**
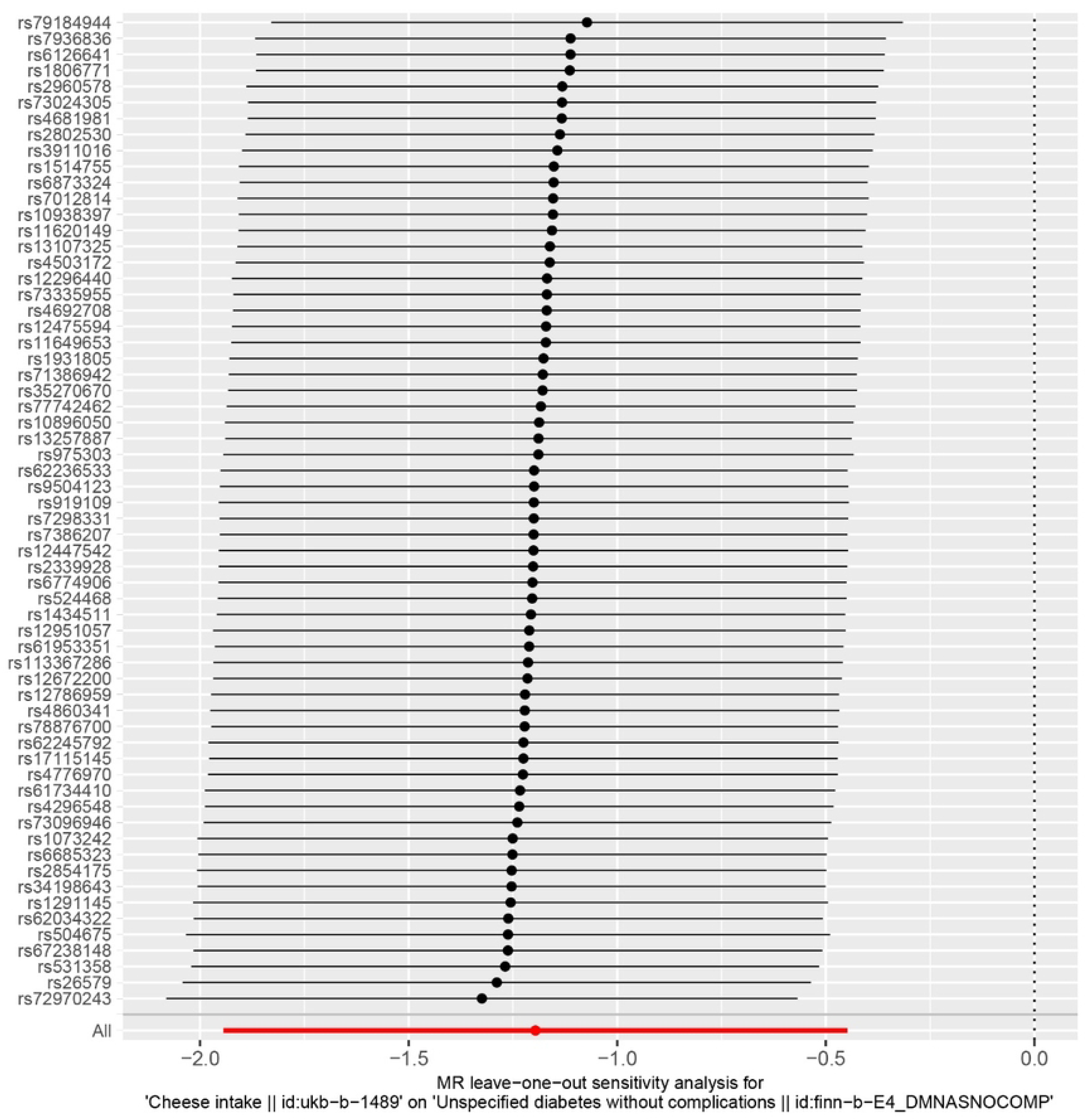
Box initially at rest on sled sliding across ice.

## 6. Discussion

With the social and economic development of countries around the world, the improvement of residents’ living standards and the change of lifestyle, the prevalence of diabetes is on the rise worldwide, becoming another chronic non communicable disease that seriously endangers public health after cardiovascular and cerebrovascular diseases and tumors^[29-31]^. Adjusting diet structure and lifestyle has become a key link in the prevention and control of diabetes. Randomized controlled trials (RCTs) are an ideal means of observing causal relationships, but due to ethical and practical limitations, the implementation of standard RCTs is relatively difficult^[32]^. Observational research can also be used to observe causal relationships, but it is susceptible to potential confounding and reverse causal relationships, making it difficult to consider as strong causal evidence. The rapid development of genetic genetics has led to the widespread application of MR in medical research. Due to the Mendelian inheritance law of random allocation during gamete formation, the association between genes and outcomes is not affected by confounding factors such as environment and acquired diseases^[33]^. MR is considered a natural RCT study. In addition, with the development of bioinformatics technology, the measurement accuracy of genetic variation continues to improve, which greatly reduces the potential bias caused by measurement errors in research. Therefore, using genetic tools as substitutes to evaluate the association between exposure and outcomes can generate more reliable causal evidence than traditional observational studies. This study found that there was a significant causal relationship between cheese intake and the onset of diabetes, and β value is less than 0 in IVW, MR Egger and weighted median methods, indicating that cheese intake is likely to be a favorable factor in preventing diabetes. The instrumental variables included in this study were all screened by the PhenoScanner database, with F value >10. The outcome data used were from two large GWAS studies, and there was no significant heterogeneity or pleiotropy between the instrumental variables. The above conditions ensure the robustness of the MR study results.

Previous research has shown that eating cheese regularly can help reduce the risk of type 2 diabetes^[34]^. Fatty foods have always been considered to increase the risk of various diseases, but the health risks associated with dairy products such as milk, butter, cheese, and yogurt are still uncertain. The new study, completed by British and Dutch researchers, involved 16800 healthy adults and 12400 patients with type 2 diabetes in eight European countries^[35]^. The conclusion of this large-scale study is that only 55 grams of cheese a day can reduce the risk of type 2 diabetes by about 12%, while other dairy products do not have the effect of cheese on reducing diabetes. Observational research has shown that consuming more dairy products (such as milk, yogurt, cheese, and whey protein) in various types of animal protein foods is beneficial for glucose regulation and/or reducing T2D risk^[36]^. Due to the conversion of carbohydrates into sugar in the body, which leads to an increase in blood sugar, T2D patients should reduce their daily carbohydrate intake. However, they can retain or even increase their ability to secrete insulin in response to protein or amino acid intake. Prospective results from adult summary analysis from the Nurse Health Study (n=72992 women), Nurse Health Study II (n=92088 women), and Health Professional Follow up Study (n=40722 men) indicate that higher plant protein intake is associated with a moderate reduction in T2DM risk^[37]^. The results also indicate that replacing animal protein foods with plant protein foods for 5% energy intake can reduce the risk of developing T2DM by about 20% -25%. When evaluating the intake of five animal protein foods (dairy products, poultry, eggs, red meat, processed meat, and fish) separately from plant protein, dairy products are the only animal protein food that is equivalent to plant protein. A group of studies on the relationship between the total intake of meat, dairy products, and fish in the population and T2DM found that as the intake of red meat and processed meat increased, the risk of developing T2D also increased by 8% and 12%, while the intake of fish and dairy products did not affect it^[38]^. Further analysis showed that the intake of high-fat fish and dairy products (cheese and fermented milk drinks) was negatively correlated with the risk of developing T2D^[39]^. A randomized equal calorie challenge study was conducted on 17 healthy adults, comparing farmhouse cheese with soy protein isolate and lean cod protein^[40]^. The results showed that compared to cod and soy protein, eating farmhouse cheese with meals resulted in a higher insulin response within four hours and a higher insulin/glucose ratio within two hours. Compared to farmhouse cheese and soy protein, the cod protein meal produced a higher glucose response within 90 minutes, but after two hours, the glucose changes in all three groups of diets tended to stabilize. These results indicate that in healthy subjects, consuming milk protein in the diet is more effective and rapid in regulating blood sugar compared to lean meat protein or soy protein. Dairy products have shown positive effects in regulating metabolism and controlling blood sugar(Figure 5). In addition to the protein factors mentioned above, other non protein dairy ingredients such as magnesium, calcium, vitamin D in fortified dairy products, fatty acids in fermented dairy products, and probiotics are all associated with a lower risk of T2DM^[41]^. Specific combinations of these compounds may have additive or synergistic effects on blood glucose regulation outcomes. The previous research results above are basically consistent with the results of this study.

**Fig. 5.**
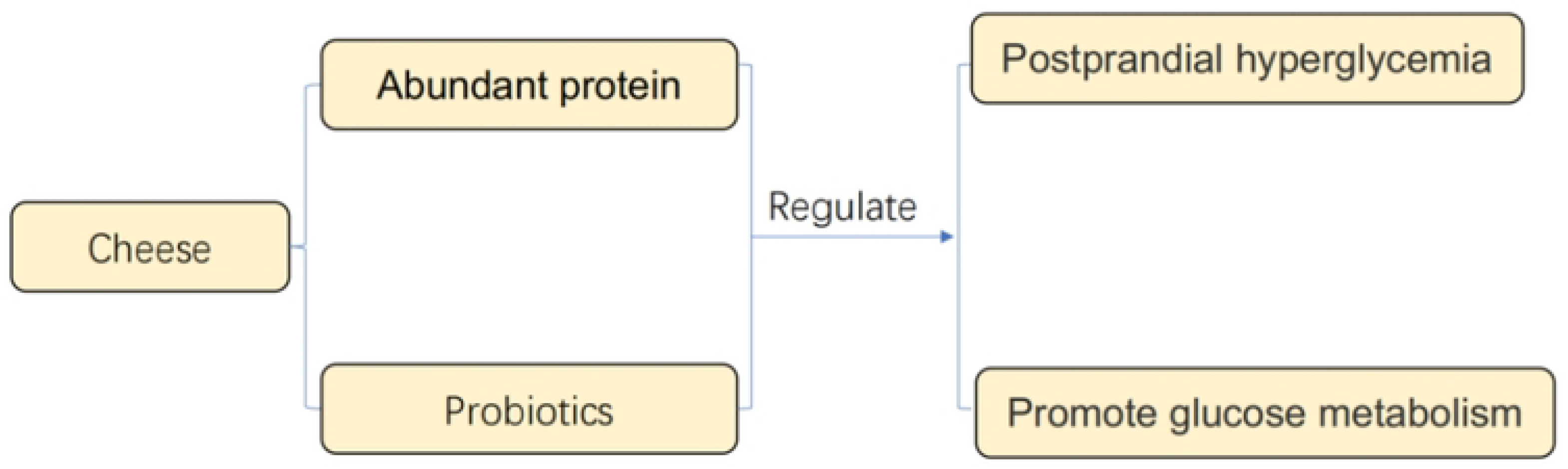
Box initially at rest on sled sliding across ice.

This study also has certain limitations. For example, this study has certain guiding significance for only focusing on the possible impact of cheese intake on the risk of diabetes or overall health, but for people who need to control blood sugar levels, the conclusions of this study are somewhat vague. A deeper focus on product characteristics (low fat, whole fat, defatted, fortified, or fermented, etc.) or protein content (such as branched chain amino acids, essential amino acids, bioactive proteins, and peptides, etc.) is a more necessary basis for evaluating and predicting glucose regulation ability.

## 7. Conclusion

The results we obtained through MR analysis support that the intake of cheese may have a causal relationship with the reduction of the risk of diabetes. In summary, in order to optimize the regulatory ability of protein intake on glucose, a comprehensive consideration should be given to the sources, types, and intake of other food drugs or supplements, as well as individual lifestyle, disease status, and actual blood glucose regulatory ability, in order to provide more personalized guidance and recommendations.

## Data Availability

If the data are held or will be held in a public repository, include URLs, accession numbers or DOIs. If this information will only be available after acceptance, indicate this by ticking the box below. For example: All XXX files are available from the XXX database (accession number(s) XXX, XXX.)

## Author Contributions

S.-Q.Y. contributed to data interpretation, the drafting of the manuscript, and approval of the article; Y. X. contributed to data interpretation and approval of the article; Y.-B.Z. contributed to critical revision of the manuscript, and approval of the article; W.-X.W. contributed to concept generation, critical revision of the manuscript, and approval of the article;

## Data Availability Statement

The data that support the findings of this study are openly available in Finnish database at http://www.finngen.fi/en and GWAS database at https://gwas.mrcieu.ac.uk.

